# Blood glucose levels in elderly subjects with type 2 diabetes during COVID-19 outbreak: a retrospective study in a single center

**DOI:** 10.1101/2020.03.31.20048579

**Authors:** Ting Xue, Qianwen Li, Qiongyao Zhang, Wei Lin, Junping Wen, Li Li, Gang Chen

## Abstract

**Aims:** Ideal glycemic control is of great importance for diabetic patients during public health emergencies of infectious diseases as long-term hyperglycemic are not only associated with chronic complications but also vital drivers of common and life-threatening infections. The present study was designed to investigate the changes of blood glucose levels in elderly subjects with type 2 diabetes(T2D) during COVID-19 outbreak.

**Methods:** This retrospective study focused on the T2D outpatients at Fujian Provincial Hospital aged 65 years old and above who received baseline test for fasting plasma glucose and/or glycated hemoglobin (HbA1c) between January 1, 2019 and March 8, 2019 and were followed up on fasting plasma glucose and/or HbA1c in the same period in 2020. The baseline and follow-up data were analyzed with the paired-samples T-test.

**Results:** A total of 135 elderly subjects with T2D with baseline and follow-up fasting plasma glucose and 50 elderly subjects with T2D with baseline and follow-up HbA1c were analyzed, respectively. The baseline and follow-up fasting plasma glucose were 7.08 ± 1.80 and 7.48±2.14 mmol/L, respectively (P=0.008). The baseline and follow-up HbA1c were 7.2±1.7% and 7.4±1.8%, respectively (P=0.158).

**Conclusions:** Elderly subjects with T2D had higher fasting plasma glucose levels during COVID-19 outbreak. We should pay more attension to the management of diabetics during public health emergencies.

## 1. Introduction

Corona virus disease 2019 (COVID-19), an infectious disease caused by severe acute respiratory syndrome coronavirus 2 (SARS-CoV-2)(aka 2019-nCoV), has widely spread all over the world since the beginning of 2020[1-3]. In the absence of antiviral drugs and vaccines to curb the epidemic, the classical public health measures were employed at an unprecedented large scale in China between January 2020 and March 2020 and gained periodical success in March 2020[4, 5]. However, with the most attention being paid to the prevention and control of the epidemic, the management of patients with chronic disease may be ignored during the response of public health emergency.

Diabetes, a chronic disease marked by glucose metabolism disorder, is one of the fastest-growing health challenges of the current society, especially in the elderly population[6]. Glycemic control is of significant importance in diabetics as complications associated with long-term hyperglycemic are not only frequent causes of disability and premature mortality but also vital drivers of indirect costs[6]. Meanwhile, the persistently elevated blood glucose levels in individuals with diabetes were considered to lead to an increase in the predisposition to infectious processes and poor prognosis[7-9]. On the one hand, hyperglycemia altered the host immune response[7-9]. Cytokines were elevated at baseline in individuals with diabetes but on stimulation were less produced than that in subjects without diabetes[7]. Dysfunction in leukocytes, lesions in monocyte and macrophage chemotaxis and phagocytosis, and damaged specific immunity have also been reported in subjects with diabetes[7-9]. On the other hand, diabetes shared several common features promoting disease progression with infectious disorders such as the pro-inflammatory state and endothelial dysfunction [9]. Both the cytokine overload in severe viral infection and the elevated cytokine synthesis in diabetes can damage the endothelium and resulted in subsequent complications[9].

Diabetes is a common underlying disease of the major infectious diseases associated with public health emergencies[10, 11]. Type 2 diabetes (T2D) accounts for the majority of all diabetes cases with high incidence in the elderly[6]. Ideal glycemic control in elderly subjects with T2D was particularly crucial during the outbreak response of public health emergencies of infectious diseases as both aging and preexisting diabetes were associated with poor prognosis of infectious diseases[7, 10, 12]. However, without antiviral drugs and vaccines in the early stage in a public health emergency of infectious disease, the classical public health measures including isolation, quarantine, social distancing, and community containment were the best and the most common approaches to curb the epidemic[4]. These approaches undoubtedly affected the lifestyles of subjects with diabetes and may have negative effect on their psychological state, which may result in blood glucose fluctuation. To our knowledge, nevertheless, no studies have explored blood glucose levels in diabetics during COVID-19 outbteak, the sixth public health emergency of international concern declared by the World Health Organization[13]. Also, no similar studies were conducted during the outbreak of previous public health emergencies of international concern those declared in 2009 for the H1N1 influenza pandemic, in 2014 for the international spread of poliovirus and the Ebola outbreak in West Africa, in 2016 for the Zika virus epidemic, and in 2019 for the Kivu Ebola epidemic[13].

The present retrospective study was performed to investigate the changes of blood glucose levels in elderly subjects with T2D during the outbreak response of COVID-19, offering the basis for the corresponding intervention strategy for the current and future public health emergencies.

## 2. Methods

### 2.1 Study Population

This retrospective study was approved by the Ethics Committee of Fujian Provincial Hospital. Due to the retrospective nature of the study, informed consent was waived. Subjects information was anonymized at the collection and analysis stage.

The present retrospective study focused on T2D outpatients at Fujian Provincial Hospital aged 65 years old and above who received baseline test for fasting plasma glucose and/or glycated hemoglobin (HbA1c) between January 1, 2019 and March 8, 2019 and were followed up on fasting plasma glucose and/or HbA1c in the same period in 2020. Subjects with conditions that affect red blood cell turnover (hemolytic and other anemias, use of drugs that stimulate erythropoiesis, glucose-6-phosphate dehydrogenase deficiency and end-stage kidney disease) were excluded. Data were obtained from the electronic health record. A total of 135 elderly subjects with T2D with baseline and follow-up fasting plasma glucose and 50 elderly subjects with T2D with baseline and follow-up HbA1c, representing elderly subjects with T2D outside Wuhan in China during the outbreak response of COVID-19, were finally analyzed, respectively.

### 2.2. Measures and definitions

The diagnosis of T2D was according to the medical records based on the International Classification of Diseases, Tenth Revision (ICD-10) codes. Fasting is defined as no caloric intake for at least 8 hours[14]. Fasting plasma glucose reflecting basal glucose levels was quantified with the hexokinase method[15]. HbA1c reflecting average glucose levels over approximately 3 months was measured by high-performance liquid chromatography[16].

### 2.3. Statistics

All statistical analyses were performed using SPSS Statistics for Windows, Version 25.0 (Armonk, NY: IBM Corp). Data are expressed as the mean ± standard deviation for continuous variables and the number(percentage) for categorical variables. Differences between the baseline and the follow-up fasting plasma glucose and that of HbA1c were both analyzed with the paired-samples T-test. All tests were two-sided and p<0.05 was defined as statistically significant.

## 3. Results

The comparison of baseline and follow-up fasting plasma glucose and that of HbA1c were shown in Table 1.

**Table 1.** The comparison of baseline and follow-up data fasting plasma glucose and that of glycated hemoglobin

|  |  | Baseline |  | Follow-up |  | P Value |
| --- | --- | --- | --- | --- | --- | --- |
| Fasting plasma glucose (n=136) | | 7.08 | $\pm 1.80$ | 7.48 | $\pm 2.14$ | 0.008* |
|  |  | mmol/L |  | mmol/L |  |  |
| Glycated hemoglobin (n=50) | | $7.2 \pm 1.7\%$ | | $7.4 \pm 1.8\%$ | | 0.158 |

Data were expressed as the mean ± standard deviation for continuous variables.
\* indicates statistical significance difference ( $p < 0.05$ ).

Among those 135 elderly subjects with T2D with baseline and follow-up fasting plasma glucose, 69(51.11%) patients were female. The follow-up fasting plasma glucose (7.48±2.14 mmol/L) was significantly higher than that of the baseline (7.08±1.80 mmol/L; p=0.008) (Table 1).

Among those 50 elderly subjects with T2D with baseline and follow-up HbA1c, 25(50.00%) patients were female. There was no statistical difference between the follow-up HbA1c (7.4±1.8%) and that of baseline (7.2±1.7%; P=0.158) (Table 1).

## 4. Discussion

The main findings of the present study were that the fasting plasma glucose levels in elderly subjects with T2D were significantly increased during the outbreak response of COVID-19. Nevertheless, statistic difference was not observed in HbA1c reflecting long-term blood glucose levels.

This is, to our knowledge, the first study to explored blood glucose levels in diabetics during the outbreak response of a public health emergency of international concern declared by the World Health Organization. With the initiation of the level-1 public health response to prevent the spread of the disease, the impacts of quarantine, social distancing and community containment during the epidemic on lifestyles may be the most important factors in the increase of fasting blood glucose in subjects with diabetes[4, 17]. Firstly, priority was given to the delivery and supply of the goods and materials needed for emergency handling of the major infection events. Although the emergency supplies and the necessities of life were guaranteed, types and amounts of fresh vegetables and fruits were less available than before. Elderly subjects with T2D had thus were forced to change their dietary associated with the good glycemic control[18]. Secondly, quarantine, social distancing and community containment always resulted in changes in physical activity, such as reduced walking and increased sedentary behaviors, which may adversely affect glycemic control in subjects with diabetes[19]. In addition, the widespread lockdown may inevitably bring an adverse psychological effect to the population, and unhealthy emotions may harm the glycemic control in subjects with diabetes[20, 21]. People with diabetes tend to have varying degrees of negative emotions, such as depression and anxiety,which may became more prominent for fear of infection during the outbreak[22, 23]. Last but not the least, to avoid cross contamination, it is inconvenient to go to hospital, especially tertiary hospitals, to buy antidiabetic drugs and glucose test strips, resulting in inadequate medication and insufficient self-monitoring of blood glucose, respectively. It’s worth noting that HbA1c was not changed during the outbreak response, indicating that plasma glucose levels rise short-term rather than long-term until March, 2020.

The results showing the changes of blood glucose levels during the outbreak response of COVID-19 were instructive for handling current and future public health emergencies. During the epidemic, most attention was paid to the prevention and control of the infection while management of chronic diseases was easy to be ignored. As mentioned above, the classical public health measures of quarantine, social distancing and community containment and its associated emotional problems may affect lifestyles and health-seeking behaviors among subjects with diabetes. However, with the infectivity and harmfulness of the major infectious diseases, especially in diabetics, home quarantine is the best choice for elderly subjects with T2D[20].

Meeting the challenge of management of elderly subjects with T2D during the outbreak response, we can focus on the following aspects to avoid the negative effects of the outbreak on diabetics. First of all, general practitioners should ont only focus on the infectious disease, but also focus on the prevention and control of chronic diseases and provide comprehensive and continuous care for elderly subjects with T2D with the context of their community. Blood glucose monitoring during the outbreak should be strengthened so that glucose levels of diabetics may return to normal as soon as possible, avoiding long-term hyperglycemia associated with chronic complications and infectious diseases[6]. To better control blood glucose levels, remote monitoring the patients’ information on blood glucose levels, exercise and diet with advanced technologies such as continuous glucose monitoring system, exercise wearables, closed-loop systems, and various smartphone applications were recommended [24, 25]. Also, telemedicine was recommended to bring convenience to the patients and make medical services more efficient[25]. In fact, telemedicine has been encouraged and promoted by the Chinese government to expand the space and content of medical services[26,27]. More and more hospitals, especially during the outbreaks, began to offer patients the service of consulting or prescribing medicine over the internet. Medicine were usually delivered by express. Patients in areas where medicine cannot be delivered by express were encouraged to bring prescriptions to the nearest pharmacy to buy the medicine after visiting the doctor online. Besides, the 5th generation wireless systems, characterized by super high data rate, low latency, high mobility, high energy efficiency, and high traffic density, should be used to develop high-quality massive open online courses (MOOCs) on health education and thus promote healthy behaviors[28]. For instance, by illustrating the nutrient composition of different foods, MOOCs on nutrition can guide diabetics to have an appropriate dietary utilizing available foods according to their physical activity and blood glucose levels. MOOCs on physical activity were also recommended to instruct the elderly subjects with T2D to do appropriate physical activity during quarantine. Health education and health-promoting behaviors have been shown to be beneficial for glycemic control[19, 29]. In addition, attention should be paid to the issue of how to maintain mental health of elderly subjects with T2D. Health workers can make use the internet, mobile applications and other online ways to carry out surveys about mental health status of diabetics, affording psychological intervention when necessary. The press workers should report the epidemic objectively and carry out proper psychological guidance to prevent spreading of negative emotion. Last but not the least, as primary care system plays an important role in the management of chronic diseases, especially during the outbreak when advanced medical care in tertiary hospitals were devoted to the battle against the major infectious disease, hierarchical medical system should be further implemented and medicine education for general practitioners should be enhanced in the future[30].

The present study has several limitations. Firstly, this is a retrospective study with a small sample size in a single center, which should not be a negligible flaw. The findings in the present study should be interpreted with some caution as the study size was quite limited. Secondly, fasting glucose was significantly affected by the diet in last night and physical activity before the test, which could not accurately reflect blood glucose control level. The results of continuous blood glucose monitoring were better indicators for short-term blood glucose levels.

## 5. Conclusions

The present study showed that the fasting plasma glucose levels were significantly increased during the outbreak response of COVID-19. Nevertheless, statistic difference was not observed in HbA1c reflecting long-term glucose level. These results indicated that we should attach more importance to the management of diabetics to help diabetes return their glucose levels to normal as soon as possible, avoiding long-term hyperglycemia associated with chronic diseases and infectious diseases during current and future outbreak.

## Data Availability

All data included in this study are available upon request by contact with the corresponding author.

## Funding

None.

## Conflict of interest

No conflict of interest has been declared by the authors

## Notes

### Competing Interest Statement

The authors have declared no competing interest.

